# Changing impact of COVID-19 vaccination on COVID-19 mortality in the SARS-CoV-2 omicron variant period, 2022-2025

**DOI:** 10.1101/2025.11.20.25340696

**Authors:** Bette Liu, Sandrine Stepien, Kristine Macartney

## Abstract

Understanding COVID-19 vaccine impact on disease outcome rates is important to inform COVID-19 vaccination program recommendations. Among 4 million Australian adults aged 65+ years we tracked COVID-19 vaccine effectiveness in preventing COVID-19-specific mortality over 7 time-periods between 2022-2025 corresponding to different SARS-CoV-2 Omicron subvariant circulation and variant-specific vaccine formulations. In January-May 2022, ancestral COVID-19 vaccine effectiveness was 93.5% (95%CI 92.6-94.2%); COVID-19 mortality rates in unvaccinated versus recently vaccine boosted individuals (within last 3 months) were 0.93 versus 0.061 per 100 person-years. By December 2024-May 2025, JN.1-variant COVID-19 relative vaccine effectiveness (compared to individuals whose last COVID-19 vaccine was >1 year ago) was 47.7% (95%CI 15.4-67.7%); COVID-19 mortality rates in unvaccinated, vaccinated >1 year ago and JN.1 booster recipients were 0.051, 0.031 and 0.016 per 100 person-years respectively. COVID-19 vaccination significantly reduced COVID-19 mortality in older adults however, the absolute mortality reduction from vaccination is less in 2025 than 2022.

## Background

As the severe acute respiratory coronavirus 2 (SARS-CoV-2) evolves, monitoring of COVID-19 vaccine effectiveness and vaccine impact on rates of specific disease outcomes is needed to inform continued vaccine programmatic and other clinical and public health decision-making. In Australia, widespread population circulation of SARS-CoV-2 began in November 2021 corresponding to opening of international borders and emergence of the highly transmissible Omicron variant^1^. Prior to Omicron circulation, high coverage (>95%)^2^ with two doses of ancestral COVID-19 vaccines was achieved in the population aged 65+ years. From 2022, at least annual boosting with ancestral and then subsequently bivalent (ancestral plus either BA.1 or BA.4/5), monovalent XBB.1.5, and most recently monovalent JN.1 subvariant COVID-19 vaccines have been recommended in this age group^3^.

Since 2022 we have documented the effectiveness of COVID-19 vaccine against COVID-19 specific mortality at various stages of the pandemic to inform immunisation policy in Australia. Published estimates included effectiveness of the ancestral vaccine^4^ (predominantly BNT162b2 mRNA Cominarty, ChAdOx-1 nCov-19 Astra Zeneca and mRNA-1273 Spikevax vaccine types), bivalent^5^ (all mRNA), and XBB.1.5 vaccines^6^ (all mRNA). These studies along with national vaccine surveillance data and reporting from other countries^7^, and the worldwide collation of COVID-19 effectiveness studies^8^, have shown that COVID-19 vaccination has been highly effective in preventing infection and severe disease, measured by hospitalisations or death, from circulating SARS-CoV-2 variants.

However vaccine effectiveness has also been shown to wane with increased time since receipt. How much of this decline in vaccine effectiveness relates to waning of an individual’s vaccine-induced specific antibody, or growing mismatch between the vaccine antigen and the circulating viral strain is difficult to measure due to correlation between both factors. This assessment is complicated further as it becomes increasingly difficult to capture the contribution of immune boosting from natural infection (in those who survive illness) over time due to reduced testing and milder and subclinical illness. Hence vaccine effectiveness measures which compare the relative rates of COVID-19 outcomes between groups based on their vaccination status can vary between studies depending on the study design, but also over time as the pandemic has progressed and the population is exposed to more waves of infection.

In this report we summarize COVID-19 vaccine effectiveness estimates and the absolute differences in COVID-19 mortality in a stable population of approximately 4 million adults living in Australia aged 65+ years over more than 3 years (2022 to mid-2025) during which time this mostly SARS-CoV-2 infection naive population became increasingly exposed^9^. We also estimated the effectiveness of the JN.1 variant vaccine against circulating JN.1-like SARS-CoV-2 variants (XEC, KP.3.1.1)^10^ in the most recent time period (December 2024-May 2025). Assessing these estimates and how they have changed over this time can inform current debate on the continuing population benefits of COVID-19 vaccine boosting in older adult populations.

## Methods

The data and detailed methods used in our assessments of COVID-19 vaccine effectiveness on mortality have been described in earlier publications^4^. Briefly, a population cohort was created using the Australian 2021 Census, which was conducted in August 2021 and includes >95% of all Australian households, linked to the Australian Immunisation Register (AIR) and national death registry. The AIR records all vaccines administered in Australia with reporting of COVID-19 vaccines mandated in February 2021. Death registrations include records of all deaths registered in Australia including the contributing causes of death coded according to the WHO International Classifications of Diseases version 10 (ICD-10). Other linked data are also available to ascertain health service use, medication use and migration. Table 1 shows each analysis period assessed, the dominant circulating SARS-CoV-2 variant based on national surveillance data, and whether estimates have been previously published. For each analysis period, we included all individuals recorded in the Census who were aged 65+ years and had not migrated from Australia or died at the beginning of that period.

**Table 1:** Seven analysis periods during which COVID-19 vaccine effectiveness was estimated, dominant circulating sub-variants, population size, COVID-19 deaths, and vaccination status, Australia 2022-2025^18,23^.

| **Analysis time period** | **Dominant circulating sub-variants** | **Population aged 65+ years\*, n** | **COVID-19 deaths** | **% unvaccinated \*** | **% more than one year since last booster\*** | **% received booster within 3 months\*** |
| --- | --- | --- | --- | --- | --- | --- |
| 1 Jan - 31 May 2022 | BA.1, BA.2 | 3,867,026 | 3,250 | 5.0 | 0.0 | 19.4 |
| 1 Jun - 30 Nov 2022 | BA.4, BA.5 | 3,877,734 | 3,185 | 4.3 | 0.0 | 34.4 |
| 1 Nov 2022 - 31 May 2023 | BA.2.75 | 4,081,257 | 2,880 | 4.2 | 0.0 | 6.2 |
| 1 Mar - 30 Sept 2023 | XB, XBB | 4,120,945 | 2,100 | 4.1 | 14.1 | 3.2 |
| 1 Aug 2023 - 28 Feb 2024 | XB, XBB, EG.5 BA.2.86 | 4,118,648 | 1,620 | 4.2 | 35.5 | 18.6 |
| 1 Mar - 30 Nov 2024 | JN.1, KP.3 | 4,223,742 | 2,435 | 4.2 | 36.0 | 17.1 |
| 1 Dec 2024 - 15 May 2025 | XEC, KP.3.1.1 | 4,264,792 | 680 | 4.1 | 50.2 | 3.6 |

\*at beginning of time period

The study outcome was COVID-19 mortality defined as a death registration where the underlying ICD-10 coded cause of death was recorded as ICD-10 U07.1 or U07.2. COVID-19 vaccine receipt was based on that recorded in the AIR. By mid-2025, vaccine recommendations meant that most older people in Australia without significant immunocompromising conditions could have received up to 8 doses of a COVID-19 vaccine, including 2 for their primary course and up to 6 boosters^21^. In all analyses we have classified individuals according to vaccination status based on recency of booster receipt. In analyses conducted in 2022 we counted the number of doses (e.g. dose 3 or dose 4) but subsequently, due to increasingly variable vaccination histories, we grouped individuals according to either having no record of vaccination, 1 or 2 doses, and then according to recency of receipt of a booster dose (>365 days [reference group], 181-365 days, 91-180 days, 8-90 days) and booster variant type if there was a newer formulation available during the time period examined.

In each analysis period individuals were followed for COVID-19 specific mortality and vaccine effectiveness was assessed using Cox proportional hazards models. Follow-up time was censored at date of death or receipt of a COVID-19 vaccine dose number one higher than the estimated maximum recommended for the general population at the time. In analyses, vaccination status was considered a time-varying covariate. Analyses were adjusted for age, sex, jurisdiction of residence, household income, number of comorbidities (assessed using an index based on pharmaceutical prescribing)^22^, number of general practice visits (as a measure of health service access) and receipt of an influenza vaccine in the year prior to the start of the time period of interest (except for household income which was based on that reported in the 2021 Census data). Adjusted hazard ratios (aHR) and 95% confidence intervals were estimated initially using unvaccinated individuals as the reference group (absolute vaccine effectiveness) but in later estimates the group whose last COVID-19 vaccine booster dose was more than 1 year previously was designated as the reference group (relative vaccine effectiveness). Vaccine effectiveness was calculated using the formula (1-aHR) * 100%. Unadjusted Quasi-Poisson regression was used to compute crude rates of COVID-19 death and death rates in vaccine status categories were estimated by multiplying the crude rate in the reference group by the aHR.

All reported counts have been perturbed using Cell Key Method, which is a technique used by the Australia Bureau of Statistics to protect the confidentiality of microdata in statistical tables by adding small, consistent amounts of noise. As all data were de-identified an exemption from ethical review was granted by the Sydney Children’s Hospital Human Research Ethics Committee.

## Results

A summary of the included study population in each of seven analysis time periods, data on COVID-19 registered deaths, and changes in the population’s vaccination uptake is provided in Table 1. The study population varied slightly in each analysis period as eligible individuals either died or migrated, new individuals entered the cohort as they turned 65 years, and due to data linkage variations. Deaths registered with the underlying cause due to COVID-19 decreased substantially over time and so did the proportion of individuals who had a recent COVID-19 vaccine booster. Characteristics of the study population according to vaccination status for each analysis has been described in the earlier publications. Those having a COVID-19 vaccine booster in the last 3 months were more likely to be older, female, have more comorbidities, more general practitioner visits and more likely to have had an influenza vaccine reported in the previous year compared to those who were less recently vaccinated.

Figure 1 shows data on the effectiveness of the JN.1 variant vaccine (available in Australia on 9 December 2024) against COVID-19 specific mortality in the latest (seventh) analysis period, relative to receipt of any vaccine dose more than a year prior in those aged 65+ years and also 75+ years. At the start of the period, 1 December 2024, as shown in Table 1, there were 4.2 million individuals of which 4.1% were unvaccinated, 50.2% had more than a year since they had last received a COVID-19 vaccine booster and 3.6% had received a COVID-19 vaccine booster in the previous 3 month period. By the analysis period end date of 15 May 2025, there were 680 COVID-19 specific deaths in the cohort. The proportion of individuals without a reported COVID-19 vaccine was similar to the beginning of the period (4.1%) but both the proportion for whom it was more than a year since vaccine receipt and the proportion who received a vaccine in the last 3 months (mostly JN.1 variant vaccine) had increased to 58.1% and 11.8% respectively. Receipt of a JN.1 vaccine, within the previous 3 months was moderately effective; the relative vaccine effectiveness was 47.7% (95%CI 15.4-67.7%) compared to vaccine receipt more than a year prior, but there was no significant benefit of other boosters (mostly XBB.1.5) given in the last 3 months nor of boosters given more than 3 months prior. Relative vaccine effectiveness was similar for those aged 75+ years as those aged 65+ years (44.7% [95%CI 10.3-65.9%]) but the absolute COVID-19 mortality rates in those aged 75+ years were about double that in those aged 65+ years. For the reference group who had received vaccine more than a year prior it was 0.062 per 100 person-years in those 75+ years compared to 0.031 per 100 person-years among all those aged 65+ years.

**Figure 1.**
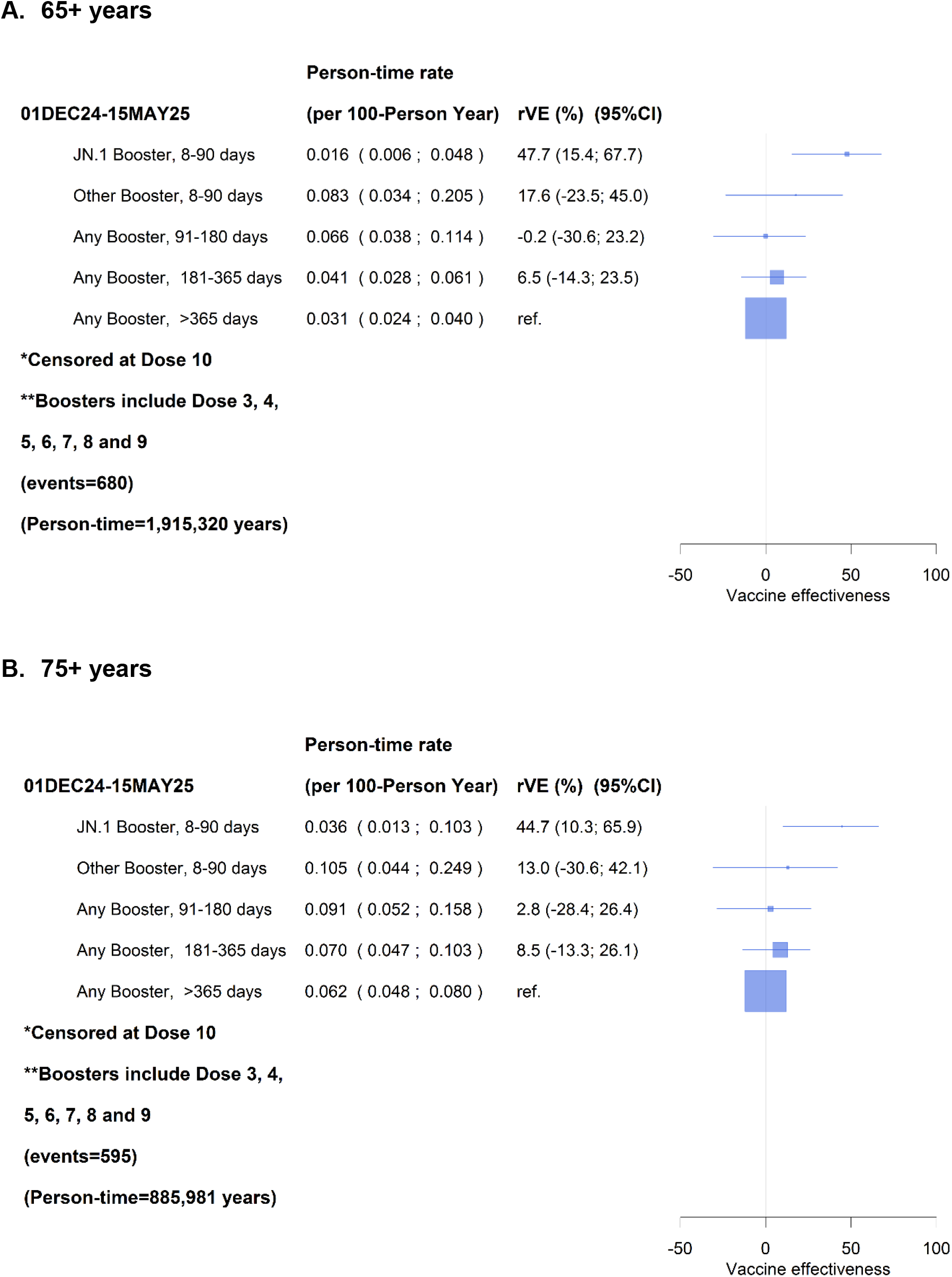
Relative effectiveness of COVID-19 vaccination against COVID-19 mortality by recency of booster and booster type, in A) 65+ years and B) 75+ years, Dec 2024-May 2025

Figure 2 shows COVID-19 vaccine effectiveness estimates against COVID-19 mortality and the estimated COVID-19 mortality rate in unvaccinated and recently (within 3 months) or less recently (>1 year) vaccine boosted individuals in all seven analysis periods between 1 January 2022 and 15 May 2025. Grouping estimates from the earlier published reports^4-6^ together with more recent unpublished effectiveness estimations enables understanding of the evolving impact of vaccination over time. Mortality rates decline substantially in all groups over the almost 3.5 years. Absolute mortality rates remained highest in the unvaccinated compared to vaccinated populations across all time periods, however, the mortality reduction in the surviving unvaccinated population over the study period was large, falling by 18-fold from 0.93 per 100 person-years in January-May 2022 to 0.051 per 100 person-years in December 2024-May 2025. For people who had received a COVID-19 booster within the last 3 months, over the same time period, there was a 3.8 fold reduction from 0.061 to 0.016 per 100 person-years. Similarly for those who had received a vaccine booster more than a year prior, there was a 3.3 fold reduction in COVID-19 mortality (from 0.103 to 0.031 per 100 person-years) over the time periods where this group was assessed (2023 onwards). These absolute differences are reflected in changes to the absolute and relative vaccine effectiveness estimates at each time period (shown in the bottom of the figure).

**Figure 2:**
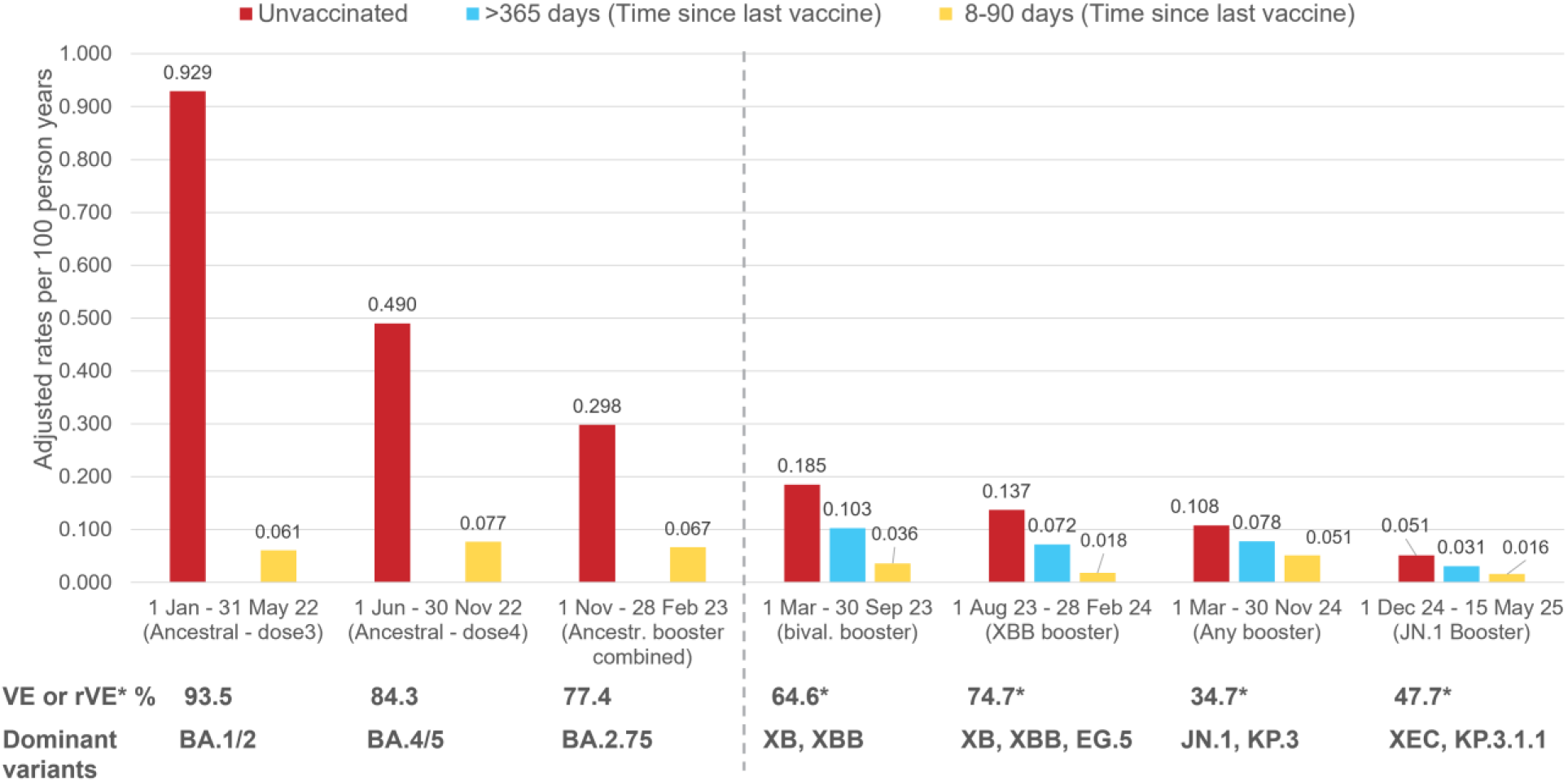
COVID-19 mortality rates* by vaccination status, 65+ years, January 2022-May 2025 Australia * Using booster more than a year ago as the reference with adjusted rates = crude rates x aHR from Cox model. Dotted line represents change from absolute vaccine effectiveness estimation to relative vaccine effectiveness.

### Interpretation

These data are the first we know of that document in the same standardised population, of almost all people residing in Australian aged 65 years and older, how COVID-19 vaccination and background SARS-CoV-2 infection have impacted on COVID-19 specific mortality over many years. Most other studies provide estimates of vaccine effectiveness only^8^ but this is a relative measure that compares disease rates between differing groups at different calendar periods. As the referent or comparison group for analyses have changed over time and varies between studies across the globe and within countries, both the interpretation as well as comparisons of estimated effects between studies, are increasingly difficult to make. Documenting both the vaccine effectiveness and the observed change in the most severe disease endpoints – in this case COVID-19 mortality rates - is an important consideration for vaccine program decision-making.

Our estimates show that in an older adult population who were mostly infection-naiive (which was the case in Australia until late 2021 due to the continent’s strict border closures), the benefits of COVID-19 vaccine boosters were very high. The relative benefits, i.e. absolute mortality reduction afforded from vaccination has fallen substantially by 2025, but remains evident in all older individuals. This is particularly the case for those older than 75 years where COVID-19 mortality is substantially higher and in whom vaccination continues to be most strongly recommended. The change observed over the years since the SAR-CoV-2 Omicron variant emerged is likely because more of the population has not only been boosted by various vaccines multiple times, but has also experienced and survived one or more SARS-CoV-2 infections, likely broadening their immunity.

We also estimated that the most recent variant-specific COVID-19 vaccine, JN.1, led to almost a halving (47.7% reduction) of COVID-19 mortality rates in adults aged 65+ years when given in the last 3 months compared to those who had not been boosted in the last year. This estimate is lower than that from a study on mortality benefit of JN.1 vaccines in Denmark^11^ and a US study on KP.2 variant vaccines on COVID-19 associated hospitalisations^12^ but similar to other studies of JN.1 variant vaccines conducted in the UK^13^ in the 3 month period following immunisation^14^. An approximate halving of COVID-19 mortality risk is a substantial reduction, and comparable to the benefit observed with influenza vaccines against serious outcomes from influenza in the same aged population, on a background of lifetime exposure to influenza viruses^15^. In the Australian population aged 65+ years during Dec 2024-May 2025 this translated to a COVID-19 death rate of 1.6/10,000 in people recently boosted with the JN.1 vaccine, compared with a rate of 3.1/10,000 deaths in those with a booster more than a year ago, and 5.1/10,000 deaths in the population with no record of vaccination. For those aged 75+ years the benefit was greater (COVID-19 death rate of 3.4/10,000 in recently boosted with the JN.1 vaccine, 6.2/10,000 in those with a booster more than a year ago). These adjusted estimates of absolute risk of COVID-19 death in 2025 provide additional utility for considering the benefits of population-based vaccination programs to the vaccine effectiveness estimate alone.

Over the study period, there were continual declines in COVID-19 mortality observed among the unvaccinated population although this mortality rate was always higher than in vaccinated populations. For the population who received a booster in the last 3 months, the greatest reductions in COVID-19 specific mortality in the recently boosted group appeared to occur in the analysis periods where the vaccine variant type matched more closely the circulating variants based on genetic phylogeny and antigenic characteristics^16,17^. These assessments were in August 2023-February 2024 when the XBB.1.5 monovalent vaccine was used for boosting and the dominant circulating viral variants included XB and XBB sub-lineages including EG.5 but also in the latter months the more distant BA.2.86^18^, and the period Dec 2024-May 2025, when the JN.1 monovalent vaccine was used and the dominant viral variants included JN.1-related lineages XEC and KP.3.1.1^19^. However other reasons for these underlying fluctuations in COVID-19 mortality in recently vaccinated individuals could also include variations in underlying COVID-19 disease rates.

Limitations of these analyses have been described earlier and include the potential for confounding by correlations between those who are more likely to get vaccinated regularly with updated COVID-19 vaccines and those with better underlying health, known as the healthy vaccinee effect. Notably though, in our data we found that those with more co-morbid conditions were more likely to get regular vaccines. Our study strengths include the use of independently collected data on vaccination and death registrations where the cause of death requires certification by a medical practitioner or coroner. Also, unlike outcomes such as hospitalisation and infection, death registration is less likely to be under-ascertained and widespread availability of free SARS-CoV-2 diagnostic testing and awareness of disease among health practitioners supports cause of death determinations. Furthermore, the use of Census data collected in 2021 and migration data means that we had a stable population where vaccination history was captured in a national register with mandated reporting by providers^20^.

Overall, our findings demonstrate the substantial benefit that COVID-19 vaccination has had on mortality reductions throughout the pandemic. However with endemicity of SARS-CoV-2 and cumulating infection-induced and hybrid immunity, as well as fewer significant evolutionary shifts in the viral variant sub-type since SARS-CoV-2 Omicron emergence in late 2021, the population-level benefits of vaccination in reducing severe disease are lower in 2025 than in earlier pandemic years. This change in the magnitude of vaccine impact over time and the waning effectiveness following vaccine administration should be considered in the decision-making for COVID-19 vaccine policy and programs, including economic estimates of cost-effectiveness, and considerations regarding who should receive COVID-19 regular vaccine boosters and the optimal timing.

## Data Availability

Data used for analyses are available on application and with appropriate ethical and governance approvals from the Australian Bureau of Statistics and data custodians.

## Data availability

Only de-identified linked data were used for these analyses. Data used for analyses are available on application and with appropriate ethical and governance approvals from the Australian Bureau of Statistics and data custodians.

## Acknowledgements

We acknowledge staff and funding from the Health Economics Research Division in the Australian Government Department of Health and Aged Care and the NHMRC Medicines Intelligence Centre for Research Excellence.

## Author contributions

BL and KM conceived the original research and managed the research. SS and BL had access to the data. SS conducted the statistical analyses. BL wrote the first draft of the paper. All authors interpreted the findings and critically reviewed and revised the manuscript.

